# Challenges in estimating waning effectiveness of two doses of BNT162b2 and ChAdOx1 COVID-19 vaccines beyond six months: an OpenSAFELY cohort study using linked electronic health records

**DOI:** 10.1101/2023.01.04.22283762

**Authors:** Elsie MF Horne, William J Hulme, Ruth H Keogh, Tom M Palmer, Elizabeth J Williamson, Edward PK Parker, Venexia M Walker, Rochelle Knight, Yinghui Wei, Kurt Taylor, Louis Fisher, Jessica Morley, Amir Mehrkar, Iain Dillingham, Sebastien Bacon, Ben Goldacre, Jonathan AC Sterne, The OpenSAFELY Collaborative

**Affiliations:** Population Health Sciences, University of Bristol, Oakfield House, Oakfield Grove, Bristol, BS8 2BN, UK; NIHR Bristol Biomedical Research Centre, Bristol, UK; Bennett Institute for Applied Data Science, Nuffield Department of Primary Care Health Sciences, University of Oxford, OX2 6GG, UK; London School of Hygiene and Tropical Medicine, Keppel Street, London, WC1E 7HT, UK; MRC Integrative Epidemiology Unit, University of Bristol, Bristol, UK; Department of Surgery, University of Pennsylvania Perelman School of Medicine, Philadelphia, PA, USA; NIHR Applied Research Collaboration West, Bristol, UK; Centre for Mathematical Sciences, School of Engineering, Computing and Mathematics, University of Plymouth, Plymouth, PL4 8AA, UK; Health Data Research UK South-West, Bristol, UK

## Abstract

Quantifying the waning effectiveness of second COVID-19 vaccination beyond six months and against the omicron variant is important for scheduling subsequent doses. With the approval of NHS England, we estimated effectiveness up to one year after second dose, but found that bias in such estimates may be substantial.

## Introduction

Understanding how effectiveness of COVID-19 vaccines changes over time and in response to new SARS-CoV-2 variants is crucial to scheduling subsequent doses. We previously quantified vaccine effectiveness (VE) over six consecutive 4-week periods from 2 to 26 weeks after second dose.^1^ Waning of hazard ratios (HRs) comparing vaccinated with unvaccinated individuals was approximately log-linear over time, and consistent across COVID-19-related outcomes and risk-based subgroups. To investigate waning beyond 26 weeks and in the omicron era, we extended follow-up to the earliest of 50 weeks after second dose or 31 March 2022.

## Methods

The data source, study design and statistical analysis were described previously.^1^ Eligible individuals were aged ≥18 years on 1 July 2020; registered at an English primary care practice using TPP SystmOne electronic health record software for ≥1 year before eligibility for COVID-19 vaccination; were not in a care home or medically housebound; and had complete demographic data with no evidence of prior SARS-CoV-2 infection.

We estimated VE across 12 consecutive 4-week comparison periods in risk-based subgroups: ages 65+, 18-64 and clinically vulnerable (CV), 40-64 and 18-39 years. We estimated VE of two doses of the BNT162b2 and ChAdOx1 vaccines (versus no vaccine) in the 65+ and 18-64 CV subgroups. VE could only be estimated for ChAdOx1 in the 40-64 subgroup, and BNT162b2 in the 18-39 subgroup.

Unvaccinated individuals were eligible for vaccination throughout follow up. From the later of September 2021 and six months after second dose, individuals at highest risk of severe COVID-19 were offered a third dose.^2,3^ Third dose eligibility was progressively extended based on risk of severe COVID-19 until mid-December 2021, when concerns about the omicron variant led to third doses being made available to all adults, with the required interval reduced to three months.^4–6^ In our analyses, unvaccinated individuals who received a first dose or vaccinated individuals who received a third dose were followed up for the remainder of that 4-week comparison period, then excluded.

## Results

There were 1,990,562, 3,281,054 and 1,227,170 eligible individuals in the BNT262b2, ChAdOx1 and unvaccinated groups respectively. Subgroup characteristics were described previously.^1^ The earliest follow-up dates in the 65+, 18-64 CV, 40-64 and 18-39 subgroups were 15 March, 21 April, 18 May and 23 July 2021 respectively. Individuals were followed for up to 50 weeks in the 65+ and 18-64 CV subgroups, and 47 and 38 weeks in the 40-64 and 18-39 subgroups respectively.

By the end of follow-up, cumulative incidence of first dose in the unvaccinated was ≤36% across all subgroups (Figure 1A). Cumulative incidence of third dose increased rapidly during the eight weeks following eligibility. In the 65+ subgroup, incidence increased from 1% 23 weeks after second dose to ≥93% by 31 weeks. Trends were similar in the 18-64 CV and 40-64 subgroups, reaching ≥90% by 50 weeks. In the 18-39 subgroup, incidence increased from 1% after 15 weeks to 62% after 23 weeks and 73% after 38 weeks.

**Figure 1.**
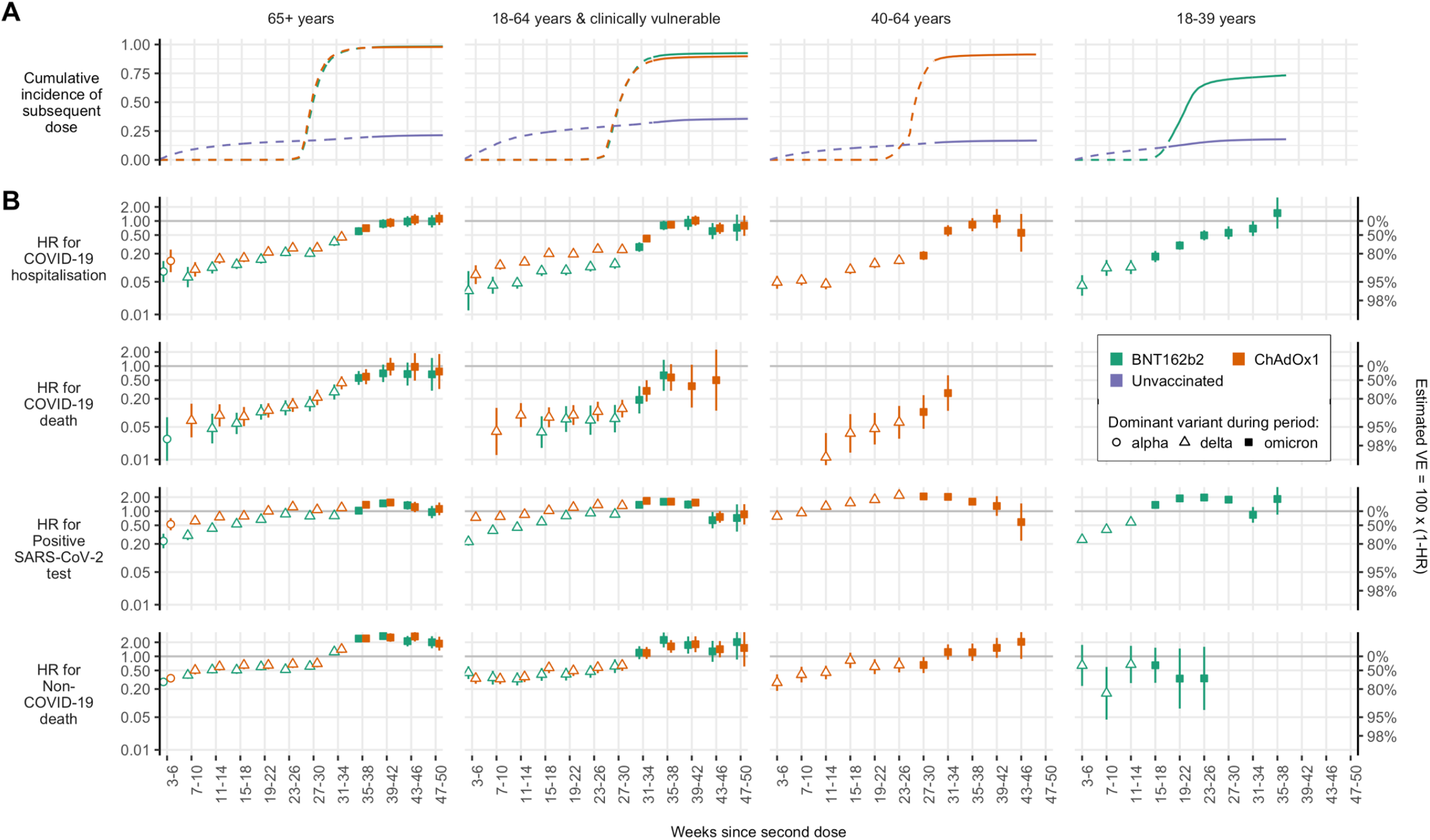
(A) Cumulative incidence of third dose in the vaccinated groups and first dose in the unvaccinated groups throughout follow-up time. Cumulative incidence lines are dashed before and solid after omicron became dominant. (B) Hazard ratios (HR) for BNT162b2 vs unvaccinated and ChAdOx1 vs unvaccinated. Shapes are hollow before and solid after omicron became dominant. Y-axes for HRs and estimated vaccine effectiveness (VE) are on the log scale. Earliest follow-up dates are as follows: 15 March 2021 in the ‘65+ years’ subgroup; 21 April 2021 in the ‘18-64 years and clinically vulnerable’ subgroup; 18 May 2021 in the ‘40-64 years’ subgroup; 23 July 2021 in the ‘18-39 years’ subgroup. The latest follow-up date in all subgroups is 31 March 2022.

Because of high uptake of third doses, estimated effectiveness of two doses during later comparison periods was based on highly selected individuals who had received two but not three doses. Estimated HRs for non-COVID-19 death in the 65+, 18-64 CV and 40-64 years subgroups changed markedly over the comparison periods during which most third doses occurred (Figure 1B). In the 65+ subgroup, estimated HRs comparing non-COVID-19 death in individuals with two BNT162b2 versus no vaccine doses increased from 0.61 (95% CI 0.51,0.73) to 2.40 (2.02,2.85) during weeks 27-30 and 35-38 respectively. Trends were similar for ChAdOx1 and the 18-64 CV and 40-64 subgroups. Because estimated HRs for non-COVID-19 death strongly suggest selection bias arising from deferred vaccination in people with a recent SARS-CoV-2 infection or in poor health, we did not attempt to interpret estimated HRs beyond 26 weeks for COVID-19-related outcomes in the 65+, 18-64 CV and 40-64 subgroups.

In the 18-39 subgroup, estimated HRs for non-COVID death (BNT162b2 only), although imprecisely estimated, did not change markedly during the rollout of third vaccine doses (Figure 1B). The cumulative incidence of third dose was lower in this than other subgroups, and postponement of vaccination because of ill-health was likely rare. Waning of HRs for COVID-19 hospitalisation was approximately log-linear over time, from 0.04 (0.03,0.07) during weeks 3-6 to 1.48 (0.69,3.17) by weeks 35-38. Waning of HRs for positive SARS-CoV-2 test was approximately log-linear up to weeks 23-26 after second dose. Estimated HRs were 0.25 (0.24,0.26) during weeks 3-6, with HRs greater than 1 by weeks 5-18. By weeks 23-26, the HR for positive SARS-CoV-2 test (1.97 (1.91,2.02)) was close to the HR for any SARS-CoV-2 test (2.16 (2.12,2.19)). HRs for any SARS-CoV-2 test remained close to 2 throughout follow-up (Supplementary Figure 2). Waning of HRs against positive SARS-CoV-2 test and COVID-19 hospitalisation in this subgroup did not appear to be affected by the emergence of the omicron variant.

## Discussion

This cohort study estimated the effectiveness of BNT162b2 and ChAdOx1 COVID-19 vaccines during 12 consecutive 4-week comparison periods, starting two weeks after second dose. Cumulative incidence of third dose in the 65+, 18-64 CV and 40-64 subgroups reached ≥90%. In these subgroups, vaccinated individuals who did not receive a third dose were at higher risk of non-COVID-19 death than unvaccinated individuals, likely due to postponement of vaccination because of recent SARS-CoV-2 infection, serious illness, or frailty. Therefore, in these subgroups estimated effectiveness of second dose against COVID-19-related outcomes is unlikely to be meaningful beyond six months, because it is based on highly selected individuals. In these subgroups, it is difficult to disentangle the effect of the omicron variant from depletion of the two-vaccine-dose group due to third dose.

In the 18-39 subgroup the maximum cumulative incidence of third dose was 73%, and there was no evidence that individuals who remained in the two-vaccine-dose group were at greater risk of non-COVID-19 death than unvaccinated individuals. Waning of HRs against COVID-19 hospitalisation in this subgroup was approximately log-linear, and VE was negligible by weeks 35-38 after second dose. Waning of HRs against positive SARS-CoV-2 test was approximately log-linear until weeks 23-26, and VE was negligible by weeks 15-18. This finding should be interpreted with caution, as it may have been due to higher uptake and reporting of SARS-CoV-2 tests in vaccinated than unvaccinated individuals. Waning HRs in the 18-39 group did not appear to be affected by emergence of the omicron variant.

An Australian survey found that unvaccinated individuals reported lower intentions both to test for SARS-CoV-2 when symptomatic and report a positive SARS-CoV-2 test than vaccinated individuals.^7^ While estimated HRs reported here were adjusted for characteristics including previously reported SARS-CoV-2 tests (Supplementary Table 1), it is likely that unmeasured confounding by testing behaviour remains. This is evident from adjusted HRs against any SARS-CoV-2 test, which were approximately 2 throughout follow-up. Waning of HRs for positive SARS-CoV-2 test in the 18-39 subgroup was approximately log-linear until weeks 23-26, then plateaued and was close to the HRs for any SARS-CoV-2 test for the remaining comparison periods (except weeks 31-34). A tentative interpretation is that estimated VE against positive SARS-CoV-2 test does not become negligible until weeks 23-26 (the inflection point in log-linear waning), while HRs ≥1 were a result of uncontrolled confounding relating to differences in testing behaviour between vaccinated and unvaccinated individuals. Follow-up from week 23 in this subgroup (Supplementary Figure 1) coincided with changes to testing policy in early January 2022,^8^ and the announcement in February that freely available mass testing would stop on 1 April 2022.^9^ End of follow-up for this study was 31 March 2022, but changes to testing behaviours are likely to have preceded this.

Third COVID-19 vaccine doses should be deferred until four weeks after the start of a SARS-CoV-2 infection.^10^ Consequently, a high proportion of individuals remaining in two-vaccine-dose groups after widespread uptake of third dose may have had current or recent SARS-CoV-2 infection. Individuals who reported a positive SARS-CoV-2 test were removed from subsequent comparison periods where the outcome was SARS-CoV-2-test-related. However, they remained in subsequent comparison periods for all other outcomes. Thus, higher prevalence of recent or current positive SARS-CoV-2 test in two-vaccine-dose groups due to delayed vaccination could have resulted in higher rates COVID-19 hospitalisation or death, and underestimates of VE against these outcomes following widespread uptake of third doses.

Previous studies reported estimates of effectiveness of second COVID-19 vaccination beyond six months^11–13^ and reported reduced VE against the omicron variant.^11^ However, the impact of third dose uptake on estimated second dose VE, and of changes in testing policy and behaviours, are rarely discussed. This study has demonstrated the importance of these factors in interpreting estimated VE. Studies increasingly focus on the incremental effectiveness of additional vaccine doses, rather than using unvaccinated individuals as the comparator. The challenges we have highlighted are also relevant to such studies. Investigators should carefully consider reasons why eligible individuals may not have received additional doses, particularly when the cumulative incidence of additional doses is high. Reporting non-COVID-19 outcomes may also provide important insights into potential biases impacting interpretation of estimated VE.

It is challenging to estimate long-term effectiveness of two COVID-19 vaccine doses in populations in which uptake of third doses was high. These challenges, which also impact investigations of VE against the omicron variant whose emergence coincided with rapid uptake of third doses, and of incremental effectiveness of third dose against second dose, should be addressed in the design and interpretation of future studies.

## Supporting information

RECORD Checklist

## Data Availability

All data were linked, stored and analysed securely within the OpenSAFELY platform: https://opensafely.org/. Data include pseudonymised data such as coded diagnoses, medications and physiological parameters. No free text data are included. All code is shared openly for review and re-use under MIT open license https://github.com/opensafely/waning-ve-2dose-1year. Detailed pseudonymised patient data is potentially re-identifiable and therefore not shared.

## Ethical Approval

This study was approved by the Health Research Authority (REC reference 22/PR/0095) and by the University of Bristol’s Faculty of Health Sciences Ethics Committee (reference 117269).

## Acknowledgements

We are very grateful for all the support received from the TPP Technical Operations team throughout this work, and for generous assistance from the information governance and database teams at NHS England and the NHS England Transformation Directorate.

