## Supplementary Materials for "Challenges in estimating waning effectiveness of two doses of BNT162b2 and ChAdOx1 COVID-19 vaccines beyond six months: an OpenSAFELY cohort study using linked electronic health records"

### Data Sharing

All data were linked, stored and analysed securely within the OpenSAFELY platform: https://opensafely.org/. Data include pseudonymised data such as coded diagnoses, medications and physiological parameters. No free text data are included. All code is shared openly for review and re-use under MIT open license https://github.com/opensafely/waning-ve-2dose-1year. Detailed pseudonymised patient data is potentially re-identifiable and therefore not shared.

Primary care records managed by the GP software provider, TPP/EMIS were linked to COVID-19 test results, hospital admissions, hospital deaths (COVID-19 only), and registered deaths through OpenSAFELY.

### Software and reproducibility

Data management was performed using Python 3.8.10, with analysis carried out using R version 4.0.2. Code for data management and analysis, as well as codelists, are archived online https://github.com/opensafely/waning-ve-2dose-1year. All iterations of the pre-specified study protocol are archived with version control https://docs.google.com/document/d/10EX8i_IfikInKRD1nkbTkLfHBJ6GdJU7PgQW_p12ibg/edit?usp=sharing.

### Information Governance

NHS England is the data controller for OpenSAFELY-TPP; TPP is the data processor; all study authors using OpenSAFELY have the approval of NHS England. This implementation of OpenSAFELY is hosted within the TPP environment which is accredited to the ISO 27001 information security standard and is NHS IG Toolkit compliant;[1]

Patient data has been pseudonymised for analysis and linkage using industry standard cryptographic hashing techniques; all pseudonymised datasets transmitted for linkage onto OpenSAFELY are encrypted; access to the platform is via a virtual private network (VPN) connection, restricted to a small group of researchers; the researchers hold contracts with NHS England and only access the platform to initiate database queries and statistical models; all database activity is logged; only aggregate statistical outputs leave the platform environment following best practice for anonymisation of results such as statistical disclosure control for low cell counts.[2]

The OpenSAFELY research platform adheres to the obligations of the UK General Data Protection Regulation (GDPR) and the Data Protection Act 2018. In March 2020, the Secretary of State for Health and Social Care used powers under the UK Health Service (Control of Patient Information) Regulations 2002 (COPI) to require organisations to process confidential patient information for the purposes of protecting public health, providing healthcare services to the public and monitoring and managing the COVID-19 outbreak and incidents of exposure; this sets aside the requirement for patient consent.[3] This was extended in July 2022 for the NHS England OpenSAFELY COVID-19 research platform.[4] In some cases of data sharing, the common law duty of confidence is met using, for example, patient consent or support from the Health Research Authority Confidentiality Advisory Group.[5]

Taken together, these provide the legal bases to link patient datasets on the OpenSAFELY platform. GP practices, from which the primary care data are obtained, are required to share relevant health information to support the public health response to the pandemic, and have been informed of the OpenSAFELY analytics platform.

3. Secretary of State for Health and Social Care. Coronavirus (COVID-19): notification to organisations to share information [Internet]. UK Government. Available from: https://web.archive.org/web/20200421171727/https://www.gov.uk/government/publications/coronavirus-covid-19-notification-of-data-controllers-to-share-information

4. Secretary of State for Health and Social Care-UK Government. Coronavirus (COVID-19): notification to organisations to share information [Internet]. Gov.uk. 2022 [cited 2022 Nov 29]. Available from: https://www.gov.uk/government/publications/coronavirus-covid-19-notification-to-organisations-to-share-information/coronavirus-covid-19-notice-under-regulation-34-of-the-health-service-control-of-patient-information-regulations-2002

5. Health Research Authority. Confidentiality Advisory Group [Internet]. NHS Health Research Authority. [cited 2022 Nov 29]. Available from: https://www.hra.nhs.uk/about-us/committees-and-services/confidentiality-advisory-group/
