## Supplementary Tables for "Challenges in estimating waning effectiveness of two doses of BNT162b2 and ChAdOx1 COVID-19 vaccines beyond six months: an OpenSAFELY cohort study using linked electronic health records"

***Supplementary Table 1: Variable definitions***

| Use | Name | Description |
|---|---|---|
| Stratification variable in all Cox models | JCVI group | Vaccine priority group defined by the UK Joint Committee on Vaccination and Immunisation (JCVI) |
| | Eligibility date | Date of eligibility for primary vaccine course |
| | Region | Region of England |
| Demographic covariate in adjusted Cox model | Age | Age on 31 March 2021 |
| | Sex | Male, Female |
| | IMD | English Index of Multiple Deprivation (IMD) quintile |
| | Ethnicity | Census categories: White, Black, South Asian, Mixed, Other |
| Clinical covariate in adjusted Cox model | BMI | Body Mass Index (BMI): Not Obese, Obese I (30-34.9 kg/m$^2$), Obese II (35-39.9 kg/m$^2$), Obese III (≥40 kg/m$^2$), |
| | Learning disability | Yes, no. |
| | Serious mental illness | Yes, no. |
| | Morbidity count | Number of morbidities in the following categories: chronic respiratory disease, chronic heart disease, chronic liver disease, chronic kidney disease, chronic neurological disease, diabetes, immunosuppression. |
| | Flu vaccine in previous 5 years | Yes, no |
| | Number of previous SARS-CoV-2 tests | Number of tests taken between 2022-05-18 and "Eligibility date": 0, 1, 2, ≥3 |
| | Pregnancy | Yes, no |

### *Supplementary Table 2: Counts and hazard ratios (HR) for COVID-19 hospitalisation the 65+ years subgroup*

| | Unvaccinated | | BNT162b2 | | | | ChAdOx1 | | | |
|---|---|---|---|---|---|---|---|---|---|---|
| weeks since second dose | n | events | n | events | unadjusted HR | adjusted HR | n | events | unadjusted HR | adjusted HR |
| 3-6 | 73,248 | 70 | 845,964 | 28 | 0.08 (0.05,0.13) | 0.08 (0.05,0.14) | 1,117,900 | 42 | 0.12 (0.08,0.20) | 0.14 (0.08,0.25) |
| 7-10 | 66,794 | 112 | 844,536 | 35 | 0.07 (0.04,0.10) | 0.06 (0.04,0.10) | 1,116,185 | 91 | 0.09 (0.07,0.13) | 0.09 (0.06,0.13) |
| 11-14 | 64,050 | 196 | 842,940 | 126 | 0.10 (0.08,0.14) | 0.10 (0.08,0.14) | 1,114,295 | 343 | 0.16 (0.13,0.19) | 0.15 (0.12,0.19) |
| 15-18 | 61,355 | 322 | 840,945 | 308 | 0.13 (0.11,0.15) | 0.12 (0.09,0.15) | 1,111,915 | 602 | 0.17 (0.14,0.20) | 0.16 (0.13,0.19) |
| 19-22 | 60,123 | 329 | 838,754 | 413 | 0.16 (0.13,0.18) | 0.15 (0.12,0.18) | 1,109,290 | 763 | 0.23 (0.20,0.27) | 0.22 (0.18,0.26) |
| 23-26 | 58,310 | 315 | 835,975 | 581 | 0.23 (0.19,0.26) | 0.21 (0.18,0.25) | 1,105,643 | 861 | 0.30 (0.25,0.35) | 0.26 (0.22,0.31) |
| 27-30 | 57,673 | 364 | 798,217 | 686 | 0.23 (0.20,0.26) | 0.20 (0.17,0.24) | 1,040,172 | 1,008 | 0.29 (0.25,0.33) | 0.26 (0.22,0.31) |
| 31-34 | 56,007 | 427 | 168,616 | 301 | 0.40 (0.34,0.48) | 0.36 (0.29,0.44) | 185,465 | 399 | 0.54 (0.46,0.63) | 0.45 (0.38,0.54) |
| 35-38 | 54,859 | 511 | 40,061 | 189 | 0.75 (0.62,0.90) | 0.60 (0.50,0.74) | 48,293 | 252 | 0.93 (0.79,1.10) | 0.70 (0.59,0.83) |
| 39-42 | 52,885 | 441 | 19,012 | 126 | 1.06 (0.86,1.31) | 0.87 (0.70,1.09) | 27,671 | 126 | 1.14 (0.92,1.43) | 0.92 (0.73,1.15) |
| 43-46 | 52,101 | 343 | 14,490 | 84 | 1.31 (1.01,1.68) | 0.97 (0.75,1.27) | 24,087 | 91 | 1.43 (1.10,1.86) | 1.07 (0.82,1.41) |
| 47-50 | 50,736 | 259 | 12,425 | 63 | 1.35 (1.01,1.81) | 0.98 (0.73,1.34) | 18,277 | 70 | 1.45 (1.07,1.97) | 1.12 (0.82,1.53) |

### *Supplementary Table 3: Counts and hazard ratios (HR) for COVID-19 death the 65+ years subgroup*

| | Unvaccinated | | BNT162b2 | | | | ChAdOx1 | | | |
|---|---|---|---|---|---|---|---|---|---|---|
| weeks since second dose | n | events | n | events | unadjusted HR | adjusted HR | n | events | unadjusted HR | adjusted HR |
| 3-6 | 73,269 | 21 | 845,992 | 7 | 0.03 (0.01,0.08) | 0.03 (0.01,0.08) | 1,117,914 | 7 | | |
| 7-10 | 66,829 | 21 | 844,585 | 7 | | | 1,116,241 | 7 | 0.05 (0.02,0.11) | 0.07 (0.03,0.16) |
| 11-14 | 64,106 | 42 | 843,017 | 14 | 0.05 (0.02,0.10) | 0.05 (0.02,0.10) | 1,114,435 | 35 | 0.10 (0.06,0.16) | 0.09 (0.05,0.15) |
| 15-18 | 61,474 | 77 | 841,127 | 42 | 0.08 (0.05,0.12) | 0.06 (0.04,0.10) | 1,112,356 | 84 | 0.10 (0.07,0.15) | 0.08 (0.05,0.13) |
| 19-22 | 60,319 | 105 | 839,195 | 98 | 0.11 (0.08,0.16) | 0.10 (0.07,0.15) | 1,110,249 | 133 | 0.12 (0.09,0.17) | 0.11 (0.08,0.16) |
| 23-26 | 58,653 | 98 | 836,731 | 112 | 0.13 (0.10,0.18) | 0.13 (0.09,0.19) | 1,107,232 | 133 | 0.17 (0.12,0.23) | 0.15 (0.10,0.21) |
| 27-30 | 58,072 | 91 | 799,386 | 133 | 0.19 (0.14,0.26) | 0.16 (0.11,0.23) | 1,042,377 | 175 | 0.25 (0.18,0.35) | 0.21 (0.15,0.31) |
| 31-34 | 56,525 | 105 | 169,659 | 77 | 0.39 (0.28,0.55) | 0.28 (0.19,0.41) | 187,117 | 119 | 0.66 (0.49,0.89) | 0.44 (0.32,0.61) |
| 35-38 | 55,496 | 126 | 40,782 | 56 | 0.79 (0.57,1.09) | 0.56 (0.40,0.79) | 49,266 | 56 | 0.84 (0.58,1.21) | 0.59 (0.41,0.86) |
| 39-42 | 53,676 | 119 | 19,481 | 35 | 0.96 (0.64,1.45) | 0.70 (0.46,1.08) | 28,378 | 35 | 1.38 (0.89,2.13) | 0.97 (0.63,1.50) |
| 43-46 | 53,060 | 98 | 14,868 | 21 | 0.98 (0.58,1.65) | 0.68 (0.38,1.19) | 24,717 | 14 | 1.23 (0.62,2.46) | 0.96 (0.49,1.89) |
| 47-50 | 51,772 | 56 | 12,782 | 14 | 0.90 (0.43,1.88) | 0.67 (0.30,1.49) | 18,816 | 7 | 0.94 (0.40,2.25) | 0.77 (0.32,1.82) |

### *Supplementary Table 4: Counts and hazard ratios (HR) for positive SARS-CoV-2 test the 65+ years subgroup*

| | Unvaccinated | | BNT162b2 | | | | ChAdOx1 | | | |
|---|---|---|---|---|---|---|---|---|---|---|
| weeks since second dose | n | events | n | events | unadjusted HR | adjusted HR | n | events | unadjusted HR | adjusted HR |
| 3-6 | 73,227 | 105 | 845,880 | 105 | 0.23 (0.16,0.31) | 0.23 (0.16,0.33) | 1,117,746 | 252 | 0.59 (0.45,0.79) | 0.53 (0.39,0.71) |
| 7-10 | 66,773 | 252 | 844,375 | 322 | 0.37 (0.30,0.45) | 0.30 (0.24,0.38) | 1,115,828 | 1,211 | 0.69 (0.59,0.81) | 0.62 (0.52,0.74) |
| 11-14 | 64,008 | 392 | 842,492 | 1,099 | 0.52 (0.46,0.60) | 0.43 (0.37,0.50) | 1,112,818 | 3,423 | 0.89 (0.79,1.00) | 0.75 (0.66,0.86) |
| 15-18 | 61,215 | 595 | 839,524 | 2,303 | 0.62 (0.56,0.69) | 0.53 (0.47,0.60) | 1,107,372 | 5,509 | 0.92 (0.84,1.02) | 0.79 (0.71,0.87) |
| 19-22 | 59,906 | 581 | 835,345 | 3,115 | 0.77 (0.69,0.85) | 0.67 (0.59,0.75) | 1,099,861 | 6,545 | 1.19 (1.08,1.31) | 1.00 (0.90,1.12) |
| 23-26 | 57,946 | 602 | 829,892 | 4,158 | 1.08 (0.98,1.19) | 0.87 (0.78,0.97) | 1,090,474 | 8,967 | 1.60 (1.46,1.76) | 1.24 (1.12,1.36) |
| 27-30 | 57,239 | 770 | 788,963 | 4,900 | 0.94 (0.86,1.02) | 0.79 (0.71,0.87) | 1,017,905 | 9,380 | 1.26 (1.16,1.37) | 1.08 (0.98,1.18) |
| 31-34 | 55,391 | 1,015 | 163,443 | 1,085 | 0.90 (0.81,1.00) | 0.80 (0.71,0.90) | 174,342 | 1,834 | 1.38 (1.26,1.52) | 1.19 (1.07,1.31) |
| 35-38 | 54,075 | 1,127 | 38,528 | 518 | 1.23 (1.10,1.39) | 1.03 (0.91,1.17) | 45,276 | 952 | 1.64 (1.48,1.80) | 1.37 (1.24,1.53) |
| 39-42 | 51,772 | 777 | 18,256 | 322 | 1.78 (1.54,2.05) | 1.47 (1.26,1.72) | 25,914 | 308 | 1.91 (1.64,2.23) | 1.54 (1.31,1.81) |
| 43-46 | 50,785 | 469 | 13,762 | 154 | 1.75 (1.43,2.14) | 1.34 (1.08,1.65) | 22,456 | 98 | 1.60 (1.24,2.07) | 1.23 (0.95,1.59) |
| 47-50 | 49,357 | 273 | 11,767 | 63 | 1.31 (0.98,1.75) | 0.96 (0.71,1.30) | 17,122 | 70 | 1.44 (1.06,1.96) | 1.12 (0.82,1.54) |

### *Supplementary Table 5: Counts and hazard ratios (HR) for non-COVID-19 death the 65+ years subgroup*

| | Unvaccinated | | BNT162b2 | | | | ChAdOx1 | | | |
|---|---|---|---|---|---|---|---|---|---|---|
| weeks since second dose | n | events | n | events | unadjusted HR | adjusted HR | n | events | unadjusted HR | adjusted HR |
| 3-6 | 73,269 | 385 | 845,992 | 854 | 0.31 (0.27,0.36) | 0.28 (0.24,0.34) | 1,117,914 | 812 | 0.43 (0.36,0.51) | 0.34 (0.28,0.41) |
| 7-10 | 66,829 | 301 | 844,585 | 1,092 | 0.44 (0.38,0.51) | 0.40 (0.33,0.47) | 1,116,241 | 973 | 0.60 (0.49,0.72) | 0.50 (0.41,0.62) |
| 11-14 | 64,106 | 280 | 843,017 | 1,274 | 0.54 (0.47,0.62) | 0.51 (0.43,0.61) | 1,114,435 | 1,148 | 0.62 (0.52,0.74) | 0.60 (0.50,0.73) |
| 15-18 | 61,474 | 259 | 841,127 | 1,337 | 0.57 (0.49,0.66) | 0.51 (0.42,0.62) | 1,112,356 | 1,253 | 0.77 (0.64,0.92) | 0.65 (0.53,0.79) |
| 19-22 | 60,319 | 259 | 839,195 | 1,393 | 0.61 (0.52,0.71) | 0.60 (0.50,0.72) | 1,110,249 | 1,190 | 0.65 (0.54,0.78) | 0.65 (0.53,0.79) |
| 23-26 | 58,653 | 273 | 836,731 | 1,568 | 0.58 (0.50,0.66) | 0.52 (0.44,0.62) | 1,107,232 | 1,372 | 0.72 (0.61,0.84) | 0.68 (0.56,0.82) |
| 27-30 | 58,072 | 280 | 799,386 | 1,582 | 0.65 (0.57,0.75) | 0.61 (0.51,0.73) | 1,042,377 | 1,400 | 0.76 (0.64,0.89) | 0.70 (0.58,0.84) |
| 31-34 | 56,525 | 266 | 169,659 | 1,057 | 1.64 (1.40,1.91) | 1.25 (1.05,1.49) | 187,117 | 791 | 1.94 (1.65,2.28) | 1.41 (1.19,1.69) |
| 35-38 | 55,496 | 287 | 40,782 | 567 | 3.33 (2.82,3.92) | 2.40 (2.02,2.85) | 49,266 | 455 | 3.40 (2.87,4.03) | 2.41 (2.03,2.87) |
| 39-42 | 53,676 | 287 | 19,481 | 266 | 3.40 (2.83,4.08) | 2.72 (2.24,3.30) | 28,378 | 203 | 3.14 (2.54,3.87) | 2.53 (2.05,3.13) |
| 43-46 | 53,060 | 252 | 14,868 | 126 | 2.63 (2.07,3.35) | 2.11 (1.63,2.73) | 24,717 | 154 | 3.31 (2.59,4.22) | 2.67 (2.08,3.42) |
| 47-50 | 51,772 | 175 | 12,782 | 91 | 2.53 (1.90,3.37) | 2.00 (1.48,2.70) | 18,816 | 70 | 2.36 (1.68,3.31) | 1.87 (1.33,2.64) |

***Supplementary Table 6: Counts and hazard ratios (HR) for Any SARS-CoV-2 test the 65+ years subgroup***

| | Unvaccinated | | BNT162b2 | | | | ChAdOx1 | | | |
|---|---|---|---|---|---|---|---|---|---|---|
| weeks since second dose | n | events | n | events | unadjusted HR | adjusted HR | n | events | unadjusted HR | adjusted HR |
| 3-6 | 73,227 | 5,810 | 845,880 | 66,556 | 2.21 (2.15,2.28) | 1.61 (1.56,1.67) | 1,117,746 | 102,060 | 2.97 (2.88,3.06) | 2.09 (2.02,2.16) |
| 7-10 | 66,773 | 5,439 | 844,375 | 74,410 | 2.55 (2.48,2.63) | 1.87 (1.80,1.93) | 1,115,828 | 113,463 | 3.12 (3.02,3.22) | 2.19 (2.12,2.27) |
| 11-14 | 64,008 | 5,516 | 842,492 | 81,816 | 2.55 (2.48,2.63) | 1.87 (1.80,1.93) | 1,112,818 | 122,906 | 3.07 (2.98,3.17) | 2.19 (2.12,2.27) |
| 15-18 | 61,215 | 5,558 | 839,524 | 81,564 | 2.48 (2.40,2.56) | 1.84 (1.78,1.91) | 1,107,372 | 120,995 | 3.00 (2.91,3.10) | 2.16 (2.09,2.23) |
| 19-22 | 59,906 | 5,537 | 835,345 | 87,339 | 2.61 (2.53,2.69) | 1.94 (1.87,2.00) | 1,099,861 | 134,113 | 3.18 (3.09,3.28) | 2.29 (2.22,2.37) |
| 23-26 | 57,946 | 5,376 | 829,892 | 96,061 | 2.87 (2.79,2.96) | 2.14 (2.07,2.21) | 1,090,474 | 146,216 | 3.45 (3.35,3.56) | 2.45 (2.38,2.53) |
| 27-30 | 57,239 | 5,551 | 788,963 | 94,430 | 2.87 (2.78,2.95) | 2.15 (2.08,2.22) | 1,017,905 | 140,140 | 3.47 (3.36,3.57) | 2.50 (2.42,2.58) |
| 31-34 | 55,391 | 5,726 | 163,443 | 18,942 | 2.41 (2.32,2.49) | 1.89 (1.82,1.97) | 174,342 | 21,119 | 2.70 (2.61,2.80) | 2.10 (2.02,2.18) |
| 35-38 | 54,075 | 5,159 | 38,528 | 5,327 | 2.85 (2.73,2.98) | 2.24 (2.13,2.35) | 45,276 | 5,852 | 3.05 (2.92,3.19) | 2.41 (2.30,2.53) |
| 39-42 | 51,772 | 4,109 | 18,256 | 2,401 | 3.31 (3.13,3.51) | 2.66 (2.50,2.83) | 25,914 | 2,576 | 3.26 (3.07,3.46) | 2.65 (2.49,2.82) |
| 43-46 | 50,785 | 3,276 | 13,762 | 1,393 | 3.27 (3.05,3.51) | 2.65 (2.46,2.85) | 22,456 | 1,701 | 3.04 (2.83,3.26) | 2.53 (2.35,2.72) |
| 47-50 | 49,357 | 2,163 | 11,767 | 931 | 3.30 (3.03,3.60) | 2.67 (2.43,2.92) | 17,122 | 1,015 | 3.15 (2.87,3.46) | 2.59 (2.36,2.85) |

***Supplementary Table 7: Counts and hazard ratios (HR) for COVID-19 hospitalisation the 18-64 years and clinically vulnerable subgroup***

| | Unvaccinated | | BNT162b2 | | | | ChAdOx1 | | | |
|---|---|---|---|---|---|---|---|---|---|---|
| weeks since second dose | n | events | n | events | unadjusted HR | adjusted HR | n | events | unadjusted HR | adjusted HR |
| 3-6 | 152,670 | 189 | 373,947 | 7 | 0.04 (0.02,0.10) | 0.03 (0.01,0.08) | 659,036 | 28 | 0.09 (0.06,0.14) | 0.07 (0.04,0.11) |
| 7-10 | 135,975 | 448 | 373,471 | 28 | 0.06 (0.04,0.08) | 0.04 (0.03,0.06) | 658,077 | 161 | 0.14 (0.12,0.17) | 0.11 (0.09,0.14) |
| 11-14 | 123,340 | 630 | 372,932 | 63 | 0.06 (0.05,0.08) | 0.05 (0.04,0.06) | 656,943 | 308 | 0.17 (0.14,0.19) | 0.13 (0.11,0.15) |
| 15-18 | 115,857 | 490 | 372,344 | 105 | 0.12 (0.10,0.15) | 0.09 (0.07,0.11) | 655,571 | 406 | 0.26 (0.23,0.30) | 0.20 (0.17,0.24) |
| 19-22 | 111,314 | 448 | 371,574 | 105 | 0.14 (0.11,0.17) | 0.09 (0.07,0.11) | 653,933 | 392 | 0.28 (0.24,0.33) | 0.20 (0.17,0.23) |
| 23-26 | 107,583 | 441 | 369,607 | 133 | 0.17 (0.14,0.21) | 0.10 (0.08,0.13) | 650,055 | 518 | 0.35 (0.30,0.40) | 0.25 (0.21,0.29) |
| 27-30 | 104,713 | 455 | 347,543 | 154 | 0.19 (0.16,0.23) | 0.12 (0.10,0.15) | 594,104 | 497 | 0.33 (0.29,0.38) | 0.24 (0.21,0.29) |
| 31-34 | 101,661 | 511 | 107,373 | 119 | 0.38 (0.31,0.47) | 0.28 (0.22,0.35) | 186,298 | 294 | 0.54 (0.46,0.63) | 0.42 (0.36,0.50) |
| 35-38 | 98,322 | 406 | 40,229 | 105 | 1.01 (0.80,1.26) | 0.80 (0.64,1.01) | 89,964 | 196 | 0.95 (0.79,1.15) | 0.83 (0.69,1.01) |
| 39-42 | 94,913 | 245 | 30,674 | 49 | 1.14 (0.80,1.62) | 0.91 (0.64,1.30) | 73,213 | 112 | 1.21 (0.95,1.54) | 1.02 (0.80,1.31) |
| 43-46 | 93,401 | 203 | 28,672 | 35 | 0.78 (0.53,1.15) | 0.61 (0.41,0.90) | 68,957 | 84 | 0.87 (0.66,1.14) | 0.70 (0.53,0.92) |
| 47-50 | 92,078 | 84 | 17,353 | 14 | 0.92 (0.50,1.71) | 0.72 (0.38,1.38) | 18,193 | 28 | 0.99 (0.62,1.60) | 0.79 (0.48,1.30) |

***Supplementary Table 8: Counts and hazard ratios (HR) for COVID-19 death the 18-64 years and clinically vulnerable subgroup***

| | Unvaccinated | | BNT162b2 | | | | ChAdOx1 | | | |
|---|---|---|---|---|---|---|---|---|---|---|
| weeks since second dose | n | events | n | events | unadjusted HR | adjusted HR | n | events | unadjusted HR | adjusted HR |
| 3-6 | 152,677 | 14 | 373,961 | 0 | | | 659,050 | 7 | | |
| 7-10 | 136,024 | 28 | 373,492 | 0 | | | 658,112 | 7 | 0.07 (0.02,0.23) | 0.04 (0.01,0.13) |
| 11-14 | 123,508 | 63 | 372,974 | 7 | | | 657,132 | 21 | 0.12 (0.07,0.20) | 0.09 (0.05,0.16) |
| 15-18 | 116,291 | 63 | 372,449 | 14 | 0.07 (0.03,0.14) | 0.04 (0.02,0.08) | 656,054 | 28 | 0.14 (0.09,0.23) | 0.08 (0.05,0.13) |
| 19-22 | 112,000 | 70 | 371,770 | 14 | 0.12 (0.07,0.23) | 0.07 (0.04,0.14) | 654,787 | 35 | 0.15 (0.10,0.24) | 0.09 (0.06,0.15) |
| 23-26 | 108,374 | 49 | 369,894 | 14 | 0.14 (0.07,0.28) | 0.07 (0.03,0.14) | 651,259 | 42 | 0.22 (0.14,0.34) | 0.11 (0.07,0.17) |
| 27-30 | 105,672 | 63 | 347,928 | 21 | 0.16 (0.09,0.28) | 0.08 (0.04,0.15) | 595,651 | 56 | 0.24 (0.16,0.36) | 0.12 (0.08,0.19) |
| 31-34 | 102,725 | 56 | 107,639 | 14 | 0.33 (0.16,0.65) | 0.19 (0.10,0.37) | 187,355 | 28 | 0.43 (0.26,0.69) | 0.29 (0.17,0.50) |
| 35-38 | 99,547 | 42 | 40,432 | 14 | 0.81 (0.38,1.73) | 0.63 (0.30,1.36) | 90,699 | 14 | 0.78 (0.41,1.48) | 0.57 (0.30,1.10) |
| 39-42 | 96,278 | 21 | 30,891 | 7 | | | 73,885 | 7 | 0.40 (0.14,1.18) | 0.37 (0.13,1.07) |
| 43-46 | 94,864 | 14 | 28,903 | 7 | | | 69,664 | 7 | 0.66 (0.18,2.45) | 0.50 (0.11,2.25) |
| 47-50 | 93,590 | 7 | 17,507 | 7 | | | 18,487 | 7 | | |

***Supplementary Table 9: Counts and hazard ratios (HR) for positive SARS-CoV-2 test the 18-64 years and clinically vulnerable subgroup***

| | Unvaccinated | | BNT162b2 | | | | ChAdOx1 | | | |
|---|---|---|---|---|---|---|---|---|---|---|
| weeks since second dose | n | events | n | events | unadjusted HR | adjusted HR | n | events | unadjusted HR | adjusted HR |
| 3-6 | 152,572 | 1,624 | 373,863 | 147 | 0.17 (0.14,0.21) | 0.22 (0.18,0.27) | 658,742 | 1,407 | 0.60 (0.55,0.65) | 0.74 (0.67,0.80) |
| 7-10 | 135,653 | 3,374 | 373,254 | 1,435 | 0.31 (0.29,0.34) | 0.39 (0.36,0.42) | 656,404 | 5,572 | 0.64 (0.61,0.67) | 0.77 (0.73,0.81) |
| 11-14 | 122,136 | 3,983 | 371,308 | 2,394 | 0.39 (0.36,0.41) | 0.45 (0.42,0.48) | 649,866 | 7,784 | 0.73 (0.70,0.76) | 0.85 (0.81,0.89) |
| 15-18 | 113,092 | 2,989 | 368,396 | 3,150 | 0.56 (0.53,0.59) | 0.59 (0.55,0.63) | 641,039 | 8,414 | 0.98 (0.93,1.02) | 1.03 (0.98,1.09) |
| 19-22 | 107,212 | 2,695 | 364,595 | 3,374 | 0.80 (0.75,0.85) | 0.80 (0.75,0.86) | 631,421 | 9,926 | 1.24 (1.19,1.30) | 1.21 (1.15,1.27) |
| 23-26 | 102,802 | 2,940 | 359,415 | 5,180 | 0.94 (0.89,0.99) | 0.92 (0.86,0.97) | 618,121 | 12,901 | 1.43 (1.37,1.50) | 1.38 (1.31,1.44) |
| 27-30 | 98,840 | 4,095 | 332,892 | 4,928 | 0.84 (0.80,0.89) | 0.86 (0.81,0.91) | 552,272 | 12,957 | 1.29 (1.24,1.35) | 1.32 (1.26,1.38) |
| 31-34 | 94,864 | 6,048 | 98,028 | 4,130 | 1.33 (1.27,1.39) | 1.37 (1.31,1.44) | 161,931 | 9,163 | 1.64 (1.58,1.70) | 1.67 (1.61,1.73) |
| 35-38 | 89,509 | 4,130 | 34,265 | 1,848 | 1.69 (1.60,1.79) | 1.59 (1.50,1.70) | 72,324 | 2,898 | 1.69 (1.60,1.78) | 1.60 (1.51,1.70) |
| 39-42 | 83,545 | 707 | 24,773 | 140 | 1.55 (1.27,1.90) | 1.39 (1.13,1.71) | 56,742 | 322 | 1.65 (1.42,1.92) | 1.53 (1.31,1.79) |
| 43-46 | 81,578 | 203 | 23,184 | 35 | 0.84 (0.57,1.25) | 0.64 (0.43,0.97) | 53,480 | 77 | 0.96 (0.73,1.27) | 0.76 (0.57,1.01) |
| 47-50 | 80,619 | 84 | 14,091 | 14 | 0.92 (0.49,1.75) | 0.72 (0.37,1.41) | 14,070 | 28 | 1.07 (0.67,1.71) | 0.86 (0.52,1.42) |

***Supplementary Table 10: Counts and hazard ratios (HR) for non-COVID-19 death the 18-64 years and clinically vulnerable subgroup***

| | Unvaccinated | | BNT162b2 | | | | ChAdOx1 | | | |
|---|---|---|---|---|---|---|---|---|---|---|
| weeks since second dose | n | events | n | events | unadjusted HR | adjusted HR | n | events | unadjusted HR | adjusted HR |
| 3-6 | 152,677 | 147 | 373,961 | 133 | 0.69 (0.53,0.91) | 0.45 (0.33,0.61) | 659,050 | 196 | 0.51 (0.40,0.64) | 0.34 (0.26,0.44) |
| 7-10 | 136,024 | 154 | 373,492 | 133 | 0.56 (0.42,0.75) | 0.35 (0.25,0.49) | 658,112 | 245 | 0.52 (0.42,0.64) | 0.33 (0.25,0.43) |
| 11-14 | 123,508 | 133 | 372,974 | 147 | 0.60 (0.46,0.79) | 0.33 (0.24,0.45) | 657,132 | 287 | 0.60 (0.48,0.75) | 0.36 (0.28,0.47) |
| 15-18 | 116,291 | 126 | 372,449 | 154 | 0.71 (0.55,0.92) | 0.41 (0.30,0.56) | 656,054 | 308 | 0.78 (0.62,0.98) | 0.57 (0.44,0.75) |
| 19-22 | 112,000 | 119 | 371,770 | 147 | 0.65 (0.49,0.86) | 0.42 (0.30,0.57) | 654,787 | 301 | 0.73 (0.57,0.93) | 0.49 (0.38,0.64) |
| 23-26 | 108,374 | 112 | 369,894 | 182 | 0.84 (0.63,1.11) | 0.47 (0.35,0.65) | 651,259 | 315 | 0.86 (0.66,1.11) | 0.58 (0.44,0.77) |
| 27-30 | 105,672 | 105 | 347,928 | 175 | 0.89 (0.68,1.17) | 0.62 (0.44,0.87) | 595,651 | 322 | 0.90 (0.71,1.14) | 0.64 (0.49,0.85) |
| 31-34 | 102,725 | 105 | 107,639 | 112 | 1.84 (1.35,2.52) | 1.20 (0.87,1.65) | 187,355 | 175 | 1.56 (1.18,2.07) | 1.18 (0.88,1.59) |
| 35-38 | 99,547 | 77 | 40,432 | 63 | 2.95 (2.06,4.24) | 2.24 (1.55,3.24) | 90,699 | 98 | 2.13 (1.56,2.93) | 1.63 (1.19,2.23) |
| 39-42 | 96,278 | 77 | 30,891 | 42 | 2.40 (1.57,3.67) | 1.75 (1.14,2.67) | 73,885 | 63 | 2.06 (1.41,2.99) | 1.82 (1.23,2.68) |
| 43-46 | 94,864 | 63 | 28,903 | 28 | 1.73 (1.05,2.83) | 1.28 (0.76,2.14) | 69,664 | 49 | 1.79 (1.23,2.62) | 1.42 (0.95,2.13) |
| 47-50 | 93,590 | 21 | 17,507 | 7 | 2.41 (0.98,5.93) | 2.02 (0.85,4.79) | 18,487 | 14 | 1.86 (0.76,4.59) | 1.51 (0.61,3.77) |

***Supplementary Table 11: Counts and hazard ratios (HR) for Any SARS-CoV-2 test the 18-64 years and clinically vulnerable subgroup***

| | Unvaccinated | | BNT162b2 | | | | ChAdOx1 | | | |
|---|---|---|---|---|---|---|---|---|---|---|
| weeks since second dose | n | events | n | events | unadjusted HR | adjusted HR | n | events | unadjusted HR | adjusted HR |
| 3-6 | 152,572 | 20,384 | 373,863 | 48,503 | 2.23 (2.19,2.27) | 2.04 (2.00,2.09) | 658,742 | 92,666 | 2.49 (2.45,2.53) | 2.23 (2.19,2.27) |
| 7-10 | 135,653 | 20,580 | 373,254 | 60,256 | 2.11 (2.07,2.14) | 1.95 (1.91,1.99) | 656,404 | 109,935 | 2.36 (2.32,2.40) | 2.11 (2.07,2.15) |
| 11-14 | 122,136 | 19,873 | 371,308 | 57,442 | 2.00 (1.96,2.03) | 1.83 (1.79,1.87) | 649,866 | 103,271 | 2.20 (2.17,2.24) | 1.98 (1.95,2.02) |
| 15-18 | 113,092 | 17,983 | 368,396 | 58,737 | 1.97 (1.93,2.01) | 1.78 (1.74,1.82) | 641,039 | 109,018 | 2.26 (2.22,2.30) | 2.01 (1.97,2.04) |
| 19-22 | 107,212 | 16,842 | 364,595 | 64,687 | 2.26 (2.22,2.30) | 2.00 (1.96,2.04) | 631,421 | 116,522 | 2.54 (2.50,2.59) | 2.20 (2.16,2.24) |
| 23-26 | 102,802 | 15,365 | 359,415 | 65,919 | 2.47 (2.42,2.52) | 2.12 (2.07,2.17) | 618,121 | 115,262 | 2.75 (2.71,2.80) | 2.35 (2.30,2.39) |
| 27-30 | 98,840 | 16,751 | 332,892 | 61,334 | 2.41 (2.37,2.46) | 2.11 (2.07,2.16) | 552,272 | 109,956 | 2.58 (2.53,2.62) | 2.24 (2.20,2.28) |
| 31-34 | 94,864 | 17,528 | 98,028 | 18,431 | 1.88 (1.84,1.92) | 1.76 (1.71,1.80) | 161,931 | 30,779 | 2.02 (1.98,2.06) | 1.90 (1.85,1.94) |
| 35-38 | 89,509 | 13,839 | 34,265 | 5,341 | 1.81 (1.75,1.88) | 1.65 (1.59,1.71) | 72,324 | 10,073 | 1.92 (1.86,1.97) | 1.74 (1.69,1.79) |
| 39-42 | 83,545 | 8,449 | 24,773 | 2,142 | 1.73 (1.65,1.82) | 1.53 (1.46,1.62) | 56,742 | 4,697 | 1.95 (1.88,2.03) | 1.75 (1.67,1.82) |
| 43-46 | 81,578 | 5,236 | 23,184 | 1,603 | 1.69 (1.59,1.79) | 1.49 (1.40,1.58) | 53,480 | 3,185 | 1.89 (1.80,1.99) | 1.69 (1.60,1.78) |
| 47-50 | 80,619 | 2,135 | 14,091 | 434 | 1.91 (1.71,2.13) | 1.74 (1.55,1.95) | 14,070 | 693 | 2.04 (1.84,2.26) | 1.81 (1.63,2.02) |

***Supplementary Table 12: Counts and hazard ratios (HR) for COVID-19 hospitalisation the 40-64 years subgroup***

| | Unvaccinated | | ChAdOx1 | | | |
|---|---|---|---|---|---|---|
| weeks since second dose | n | events | n | events | unadjusted HR | adjusted HR |
| 3-6 | 315,294 | 511 | 1,504,055 | 63 | 0.06 (0.04,0.08) | 0.05 (0.04,0.07) |
| 7-10 | 299,026 | 588 | 1,502,354 | 112 | 0.06 (0.05,0.07) | 0.05 (0.04,0.07) |
| 11-14 | 288,365 | 602 | 1,500,534 | 126 | 0.05 (0.04,0.07) | 0.04 (0.03,0.06) |
| 15-18 | 280,805 | 525 | 1,498,819 | 175 | 0.10 (0.09,0.13) | 0.09 (0.07,0.11) |
| 19-22 | 275,310 | 504 | 1,496,586 | 210 | 0.14 (0.12,0.17) | 0.12 (0.10,0.15) |
| 23-26 | 270,340 | 553 | 1,468,761 | 322 | 0.18 (0.15,0.21) | 0.14 (0.12,0.17) |
| 27-30 | 265,125 | 497 | 1,026,424 | 273 | 0.19 (0.16,0.23) | 0.18 (0.15,0.23) |
| 31-34 | 259,210 | 273 | 206,864 | 119 | 0.74 (0.58,0.95) | 0.62 (0.47,0.81) |
| 35-38 | 255,199 | 154 | 148,925 | 63 | 1.02 (0.76,1.39) | 0.83 (0.61,1.15) |
| 39-42 | 252,147 | 70 | 113,092 | 42 | 1.48 (0.96,2.28) | 1.12 (0.70,1.80) |
| 43-46 | 99,624 | 28 | 59,332 | 7 | 0.76 (0.34,1.69) | 0.56 (0.22,1.41) |
| 47-50 | 3,423 | 0 | 70 | 0 | | |

***Supplementary Table 13: Counts and hazard ratios (HR) for COVID-19 death the 40-64 years subgroup***

| | Unvaccinated | | ChAdOx1 | | | |
|---|---|---|---|---|---|---|
| weeks since second dose | n | events | n | events | unadjusted HR | adjusted HR |
| 3-6 | 315,329 | 14 | 1,504,090 | 7 | | |
| 7-10 | 299,250 | 21 | 1,502,452 | 7 | | |
| 11-14 | 288,862 | 49 | 1,500,744 | 7 | 0.02 (0.01,0.05) | 0.01 (0.00,0.04) |
| 15-18 | 281,512 | 35 | 1,499,148 | 7 | 0.04 (0.02,0.09) | 0.04 (0.01,0.09) |
| 19-22 | 276,241 | 35 | 1,497,076 | 14 | 0.06 (0.03,0.13) | 0.05 (0.02,0.10) |
| 23-26 | 271,425 | 42 | 1,469,447 | 21 | 0.07 (0.04,0.14) | 0.06 (0.03,0.14) |
| 27-30 | 266,399 | 42 | 1,027,222 | 21 | 0.14 (0.07,0.28) | 0.10 (0.05,0.24) |
| 31-34 | 260,659 | 35 | 207,333 | 7 | 0.29 (0.12,0.66) | 0.26 (0.11,0.64) |
| 35-38 | 256,767 | 14 | 149,324 | 7 | | |
| 39-42 | 253,722 | 7 | 113,442 | 7 | | |
| 43-46 | 100,436 | 7 | 59,577 | 0 | | |
| 47-50 | 3,465 | 0 | 70 | 0 | | |

***Supplementary Table 14: Counts and hazard ratios (HR) for positive SARS-CoV-2 test the 40-64 years subgroup***

| | Unvaccinated | | ChAdOx1 | | | |
|---|---|---|---|---|---|---|
| weeks since second dose | n | events | n | events | unadjusted HR | adjusted HR |
| 3-6 | 314,615 | 5,866 | 1,500,366 | 9,100 | 0.91 (0.88,0.95) | 0.78 (0.75,0.81) |
| 7-10 | 296,149 | 5,985 | 1,489,642 | 13,692 | 1.06 (1.02,1.09) | 0.92 (0.89,0.96) |
| 11-14 | 282,863 | 5,670 | 1,474,256 | 17,731 | 1.48 (1.43,1.54) | 1.27 (1.22,1.32) |
| 15-18 | 273,161 | 5,887 | 1,454,971 | 20,734 | 1.83 (1.77,1.89) | 1.52 (1.46,1.57) |
| 19-22 | 265,405 | 6,860 | 1,432,242 | 29,239 | 2.17 (2.10,2.23) | 1.77 (1.71,1.83) |
| 23-26 | 258,083 | 9,884 | 1,377,586 | 43,428 | 2.60 (2.54,2.67) | 2.18 (2.13,2.24) |
| 27-30 | 249,795 | 9,632 | 934,052 | 33,656 | 2.37 (2.30,2.44) | 2.07 (2.01,2.14) |
| 31-34 | 238,924 | 4,417 | 160,083 | 5,320 | 2.35 (2.24,2.46) | 2.03 (1.93,2.13) |
| 35-38 | 231,945 | 861 | 114,191 | 322 | 1.95 (1.69,2.25) | 1.60 (1.38,1.86) |
| 39-42 | 228,557 | 70 | 89,033 | 42 | 1.71 (1.09,2.69) | 1.29 (0.80,2.10) |
| 43-46 | 92,120 | 28 | 48,832 | 7 | 0.80 (0.36,1.79) | 0.59 (0.23,1.47) |
| 47-50 | 3,255 | 0 | 63 | 0 | | |

***Supplementary Table 15: Counts and hazard ratios (HR) for non-COVID-19 death the 40-64 years subgroup***

| | Unvaccinated | | ChAdOx1 | | | |
|---|---|---|---|---|---|---|
| weeks since second dose | n | events | n | events | unadjusted HR | adjusted HR |
| 3-6 | 315,329 | 98 | 1,504,090 | 91 | 0.25 (0.18,0.35) | 0.27 (0.18,0.41) |
| 7-10 | 299,250 | 77 | 1,502,452 | 119 | 0.39 (0.28,0.54) | 0.41 (0.28,0.60) |
| 11-14 | 288,862 | 84 | 1,500,744 | 126 | 0.41 (0.30,0.56) | 0.45 (0.32,0.63) |
| 15-18 | 281,512 | 63 | 1,499,148 | 161 | 0.75 (0.53,1.06) | 0.82 (0.56,1.20) |
| 19-22 | 276,241 | 77 | 1,497,076 | 168 | 0.57 (0.41,0.79) | 0.60 (0.42,0.87) |
| 23-26 | 271,425 | 70 | 1,469,447 | 168 | 0.56 (0.40,0.78) | 0.66 (0.45,0.95) |
| 27-30 | 266,399 | 70 | 1,027,222 | 140 | 0.58 (0.40,0.83) | 0.65 (0.44,0.97) |
| 31-34 | 260,659 | 70 | 207,333 | 63 | 1.33 (0.91,1.95) | 1.23 (0.84,1.80) |
| 35-38 | 256,767 | 63 | 149,324 | 42 | 1.41 (0.94,2.12) | 1.22 (0.80,1.87) |
| 39-42 | 253,722 | 42 | 113,442 | 35 | 1.60 (0.99,2.60) | 1.52 (0.93,2.48) |
| 43-46 | 100,436 | 21 | 59,577 | 14 | 2.13 (0.93,4.91) | 2.05 (0.88,4.74) |
| 47-50 | 3,465 | 0 | 70 | 0 | | |

***Supplementary Table 16: Counts and hazard ratios (HR) for Any SARS-CoV-2 test the 40-64 years subgroup***

| | Unvaccinated | | ChAdOx1 | | | |
|---|---|---|---|---|---|---|
| weeks since second dose | n | events | n | events | unadjusted HR | adjusted HR |
| 3-6 | 314,615 | 34,706 | 1,500,366 | 246,358 | 3.81 (3.76,3.86) | 2.83 (2.79,2.87) |
| 7-10 | 296,149 | 34,566 | 1,489,642 | 254,555 | 3.53 (3.48,3.57) | 2.67 (2.64,2.71) |
| 11-14 | 282,863 | 32,788 | 1,474,256 | 255,353 | 3.75 (3.70,3.80) | 2.86 (2.82,2.90) |
| 15-18 | 273,161 | 30,786 | 1,454,971 | 279,734 | 4.15 (4.10,4.21) | 3.15 (3.11,3.19) |
| 19-22 | 265,405 | 31,465 | 1,432,242 | 289,226 | 4.28 (4.23,4.34) | 3.26 (3.21,3.30) |
| 23-26 | 258,083 | 33,460 | 1,377,586 | 296,072 | 4.42 (4.37,4.48) | 3.40 (3.35,3.44) |
| 27-30 | 249,795 | 30,898 | 934,052 | 211,099 | 3.73 (3.68,3.79) | 2.89 (2.84,2.93) |
| 31-34 | 238,924 | 21,350 | 160,083 | 19,117 | 2.68 (2.62,2.74) | 2.21 (2.16,2.26) |
| 35-38 | 231,945 | 13,125 | 114,191 | 7,343 | 2.31 (2.24,2.39) | 1.90 (1.83,1.96) |
| 39-42 | 228,557 | 7,266 | 89,033 | 4,340 | 2.49 (2.39,2.61) | 2.05 (1.95,2.14) |
| 43-46 | 92,120 | 2,541 | 48,832 | 1,407 | 2.84 (2.65,3.06) | 2.28 (2.11,2.46) |
| 47-50 | 3,255 | 28 | 63 | 7 | | |

***Supplementary Table 17: Counts and hazard ratios (HR) for COVID-19 hospitalisation the 18-39 years subgroup***

| | Unvaccinated | | BNT162b2 | | | |
|---|---|---|---|---|---|---|
| weeks since second dose | n | events | n | events | unadjusted HR | adjusted HR |
| 3-6 | 685,622 | 763 | 706,454 | 21 | 0.04 (0.02,0.06) | 0.04 (0.03,0.07) |
| 7-10 | 653,359 | 560 | 701,974 | 28 | 0.09 (0.06,0.14) | 0.10 (0.07,0.15) |
| 11-14 | 630,420 | 574 | 698,698 | 42 | 0.11 (0.08,0.15) | 0.10 (0.07,0.15) |
| 15-18 | 613,081 | 665 | 695,541 | 70 | 0.18 (0.14,0.23) | 0.17 (0.13,0.23) |
| 19-22 | 595,175 | 791 | 584,339 | 126 | 0.28 (0.23,0.34) | 0.30 (0.24,0.37) |
| 23-26 | 574,707 | 630 | 305,249 | 84 | 0.50 (0.40,0.64) | 0.49 (0.38,0.63) |
| 27-30 | 557,340 | 392 | 219,170 | 56 | 0.63 (0.46,0.85) | 0.56 (0.41,0.77) |
| 31-34 | 548,149 | 224 | 178,864 | 42 | 0.89 (0.63,1.26) | 0.69 (0.48,0.99) |
| 35-38 | 350,091 | 56 | 52,241 | 14 | 1.97 (0.96,4.05) | 1.48 (0.69,3.17) |

***Supplementary Table 18: Counts and hazard ratios (HR) for positive SARS-CoV-2 test the 18-39 years subgroup***

| | Unvaccinated | | BNT162b2 | | | |
|---|---|---|---|---|---|---|
| weeks since second dose | n | events | n | events | unadjusted HR | adjusted HR |
| 3-6 | 678,657 | 16,450 | 701,554 | 2,744 | 0.31 (0.30,0.32) | 0.25 (0.24,0.26) |
| 7-10 | 637,630 | 14,203 | 694,400 | 4,151 | 0.52 (0.50,0.54) | 0.41 (0.39,0.42) |
| 11-14 | 609,763 | 16,688 | 687,043 | 7,420 | 0.75 (0.73,0.78) | 0.58 (0.56,0.60) |
| 15-18 | 586,173 | 29,519 | 676,578 | 27,195 | 1.71 (1.68,1.74) | 1.36 (1.34,1.39) |
| 19-22 | 561,239 | 35,399 | 543,718 | 42,980 | 2.32 (2.29,2.36) | 1.88 (1.85,1.91) |
| 23-26 | 523,810 | 18,200 | 245,630 | 12,880 | 2.52 (2.46,2.58) | 1.97 (1.91,2.02) |
| 27-30 | 494,970 | 3,325 | 164,507 | 707 | 2.38 (2.18,2.59) | 1.77 (1.62,1.94) |
| 31-34 | 484,484 | 196 | 134,981 | 42 | 1.11 (0.77,1.60) | 0.84 (0.57,1.23) |
| 35-38 | 310,051 | 56 | 38,731 | 14 | 2.54 (1.23,5.23) | 1.83 (0.85,3.94) |

***Supplementary Table 19: Counts and hazard ratios (HR) for non-COVID-19 death the 18-39 years subgroup***

| | Unvaccinated | | BNT162b2 | | | |
|---|---|---|---|---|---|---|
| weeks since second dose | n | events | n | events | unadjusted HR | adjusted HR |
| 3-6 | 685,895 | 28 | 706,468 | 7 | 0.66 (0.27,1.59) | 0.64 (0.23,1.76) |
| 7-10 | 654,024 | 28 | 702,009 | 7 | 0.18 (0.05,0.64) | 0.16 (0.04,0.59) |
| 11-14 | 631,344 | 21 | 698,761 | 7 | 0.57 (0.21,1.53) | 0.67 (0.27,1.68) |
| 15-18 | 614,145 | 21 | 695,639 | 14 | 0.65 (0.29,1.45) | 0.64 (0.27,1.53) |
| 19-22 | 596,470 | 21 | 584,493 | 7 | 0.36 (0.09,1.48) | 0.34 (0.08,1.47) |
| 23-26 | 576,205 | 21 | 305,452 | 7 | 0.41 (0.10,1.71) | 0.34 (0.07,1.60) |
| 27-30 | 559,132 | 14 | 219,408 | 7 | | |
| 31-34 | 550,060 | 7 | 179,095 | 7 | | |
| 35-38 | 351,512 | 7 | 52,325 | 0 | | |

***Supplementary Table 20: Counts and hazard ratios (HR) for Any SARS-CoV-2 test the 18-39 years subgroup***

| | Unvaccinated | | BNT162b2 | | | |
|---|---|---|---|---|---|---|
| weeks since second dose | n | events | n | events | unadjusted HR | adjusted HR |
| 3-6 | 678,657 | 101,213 | 701,554 | 119,784 | 2.35 (2.32,2.37) | 1.82 (1.80,1.84) |
| 7-10 | 637,630 | 86,037 | 694,400 | 120,169 | 2.71 (2.69,2.74) | 2.09 (2.07,2.11) |
| 11-14 | 609,763 | 81,767 | 687,043 | 117,593 | 2.81 (2.78,2.84) | 2.13 (2.11,2.15) |
| 15-18 | 586,173 | 100,380 | 676,578 | 152,446 | 2.76 (2.73,2.78) | 2.10 (2.08,2.12) |
| 19-22 | 561,239 | 100,191 | 543,718 | 145,754 | 3.02 (2.99,3.04) | 2.30 (2.27,2.32) |
| 23-26 | 523,810 | 63,567 | 245,630 | 41,895 | 2.83 (2.79,2.87) | 2.16 (2.12,2.19) |
| 27-30 | 494,970 | 34,517 | 164,507 | 12,439 | 2.43 (2.38,2.48) | 1.88 (1.84,1.93) |
| 31-34 | 484,484 | 18,284 | 134,981 | 6,174 | 2.39 (2.32,2.47) | 1.84 (1.78,1.90) |
| 35-38 | 310,051 | 5,299 | 38,731 | 1,141 | 2.68 (2.51,2.87) | 1.92 (1.79,2.06) |
