## Supplementary Figures for "Challenges in estimating waning effectiveness of two doses of BNT162b2 and ChAdOx1 COVID-19 vaccines beyond six months: an OpenSAFELY cohort study using linked electronic health records"

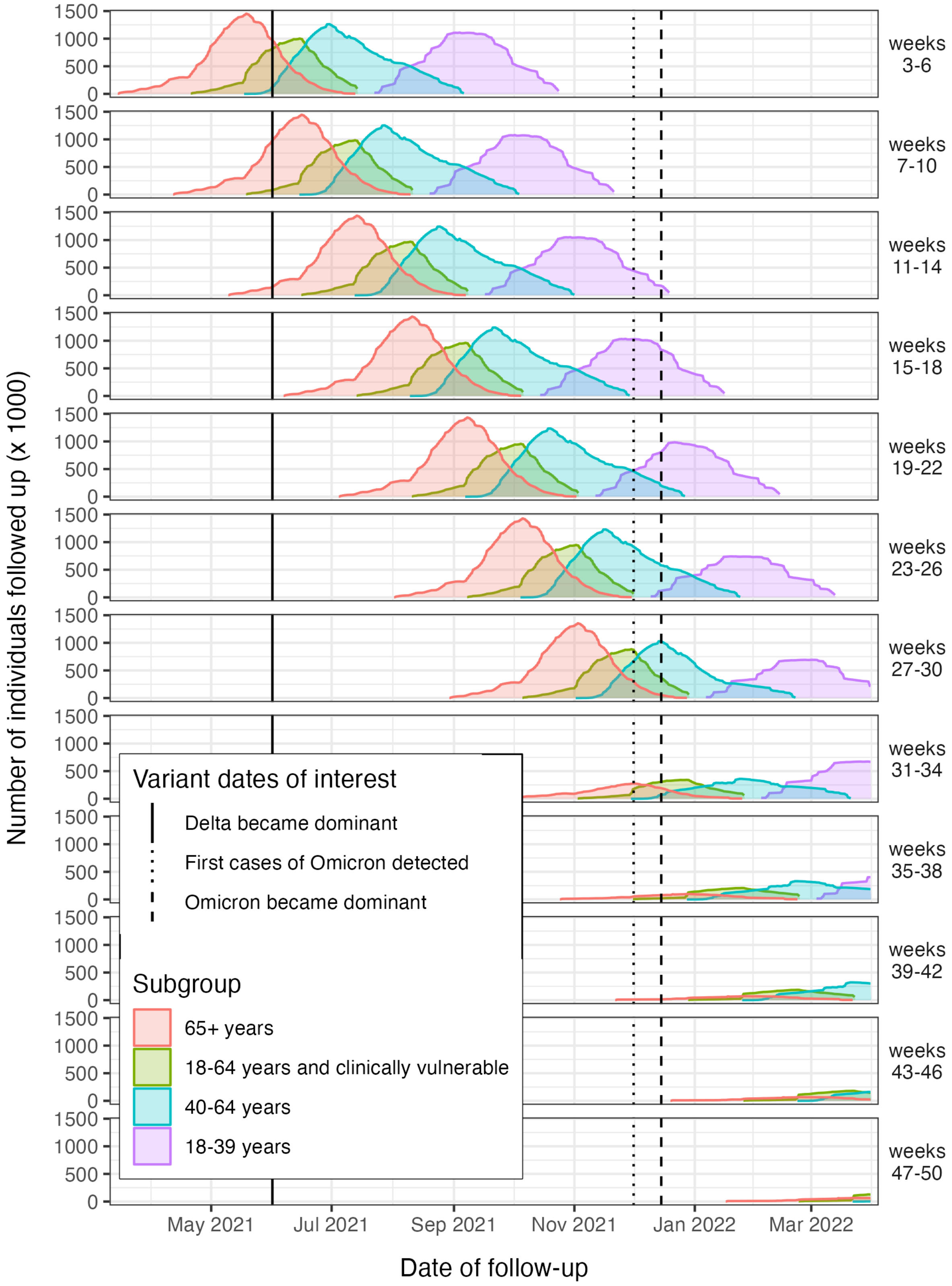


*Supplementary Figure 1. Distribution of follow-up time across comparison periods (defined by weeks since second dose)*

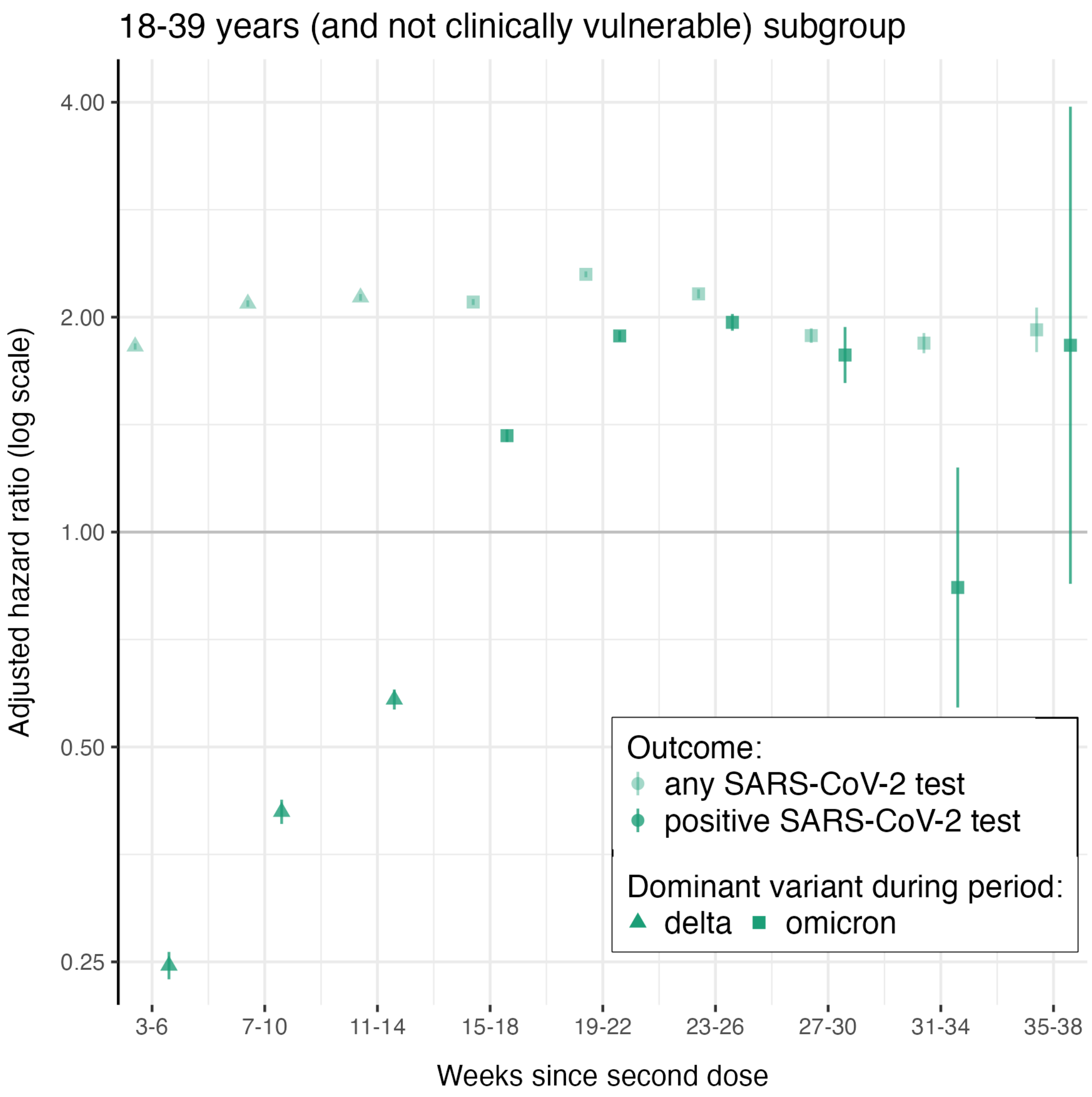


*Supplementary Figure 2. Hazard ratios for SARS-CoV-2 test and positive SARS-CoV-2 test in the 18-39 years subgroup. Follow-up was censored for both outcomes at the date of the first positive SARS-CoV-2 test. Only the first SARS-CoV-2 test in each comparison period (defined by weeks since second dose) was counted. The calendar time corresponding to each comparison period can be inferred from Supplementary Figure 1.*
